# The effectiveness of precision supplements on depression symptoms in a US population

**DOI:** 10.1101/2023.04.18.23288750

**Authors:** Cristina Julian, Nan Shen, Matthew Molusky, Lan Hu, Vishakh Gopu, Anmol Gorakshakar, Eric Patridge, Grant Antoine, Janelle Connell, Hilary Keiser, Uma Naidoo, Momchilo Vuyisich, Guruduth Banavar

## Abstract

Nutrition plays a pivotal role in depression, but dietary interventions are usually not personalized and do not consider patients’ microbial and human molecular functions. This preliminary study evaluated the effectiveness of precision supplements (PS) on depression symptoms as part of a personalized nutrition subscription plan that accounts for the gene expression activity of the microbiome and the human host. People with depression, 86 taking PS and 45 controls responded to the patient health questionnaire-9 (PHQ-9) at two time points an average of ∼6 months apart. Categorical changes were evaluated using the PHQ-9 score system, and clinically significant categorical differences were observed between the two groups (effect size = 0.48; p <0.001). The difference in differences was calculated using multiple group propensity score weighting adjusting for age, sex, BMI, and physical activity, and the PHQ-9 score decreased by ∼4 points (∼29%) for the intervention group (t0: 13.75+-3.80, t1: 9.78+-6.42) vs Controls (t0: 14.07+-3.64, t1: 13.59 +-6.65). Thus, precision supplement use over ∼6 months significantly reduced depression symptoms, with 69.8% of the individuals in the intervention group improving their category to no/low depression vs. 15.6% in the control group.

## 1. Introduction

Depression is a common, but potentially lethal mood disorder, a leading cause of disability worldwide. Symptoms can include feelings of hopelessness, irritability, loss of interest in activities, fatigue, difficulty concentrating or sleeping, and even thoughts of death or suicidal ideation. In 2020, an estimated 21 million adults in the United States had at least one major depressive episode, representing 8.4% of all US adults [1]. The rate of serious mental health issues such as depression have more than doubled in the USA during the pandemic [2].

Depression is usually treated with medications, psychotherapy, or a combination of the two. More severe cases may need treatments such as electroconvulsive therapy (ECT) or rTMS (repetitive transcranial magnetic stimulation). Unfortunately, about 30% to 40% of patients do not adequately respond to pharmacotherapy and other therapies [3], which makes it difficult to improve the mood disorder or the patient’s quality of life. There is growing evidence that diet and nutrition play a pivotal role in maintaining good mental health. Specific ingredients like probiotics, micronutrients, omega-3 polyunsaturated fatty acids, dietary fibers, and polyphenols have shown antidepressant effects in various ways. For example, a health survey in women showed that the symptoms of depression worsened as the intake of dietary fiber decreased [4]. A recent meta-analysis concluded that it is likely that daily consumption of a probiotic supplement could have a positive effect in improving the mood, anxiety, and cognitive symptoms present in major depression [5]. People with depression seem to have lower levels of vitamin B9 [6], and vitamin B complex is essential for neuronal function, and deficiencies have been linked to depression [7]. Vitamin D has been associated with reducing the development and symptoms of depression [8], and antioxidants have therapeutic potential as adjunctive therapy to conventional antidepressants [9]. While several single supplement ingredients have been associated with the treatment of depression, the role of precision supplements including several ingredients based on an individual’s overall condition has not been addressed.

Viome is a wellness company that identifies early signs of microbial and human imbalance and inflammation in the body by profiling the stool, saliva and blood samples packaged in different kit options, processed in the lab using a metatranscriptomic approach[10]. The Viome service provides personalized food and supplement recommendations based on an algorithm that takes into account the gut and human biochemical functions and self-reported phenotype information [11]. The algorithm computes a set of ingredients with dosages targeting all Viome-assigned conditions for that individual. These ingredients subsequently are made available to individuals through precision supplements. In addition, the Viome platform includes clinically validated questionnaires where individuals can report the signs and symptoms of a particular disease. In particular, the Patient Health Questionnaire-9 (PHQ-9) [12] has been used since 2019 to evaluate symptoms of depression.

Since we are evaluating under real-world conditions [13], the primary goal of this study was to assess the effectiveness of Viome precision supplements on depression symptoms as measured by PHQ-9.

## 2. Materials and Methods

### 2.1. Study population

This study used data from the Viome database up to 1 December 2022 (Figure 1). Eligible participants were defined as those Viome customers who purchased at least two kits, with results and recommendations released, who were aged ≥18 years at the time of purchasing their first Viome kit, and who responded to the patient health questionnaire (PHQ-9) at the time of kit purchase. All customers consented to their data being used for research purposes, as part of the sign-up process for the Viome services. A federally accredited Institutional Review Board in the USA approved the study.

**Figure 1.**
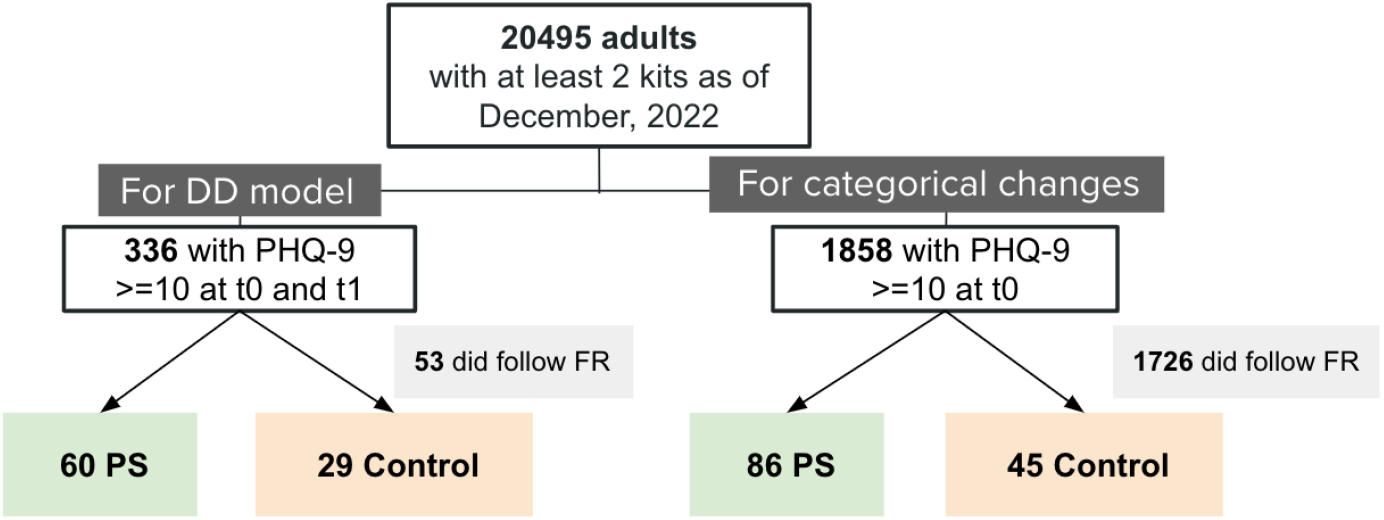
Study population flowchart FR, food recommendations; DD, difference in difference, PHQ-9, patient health questionnaire 9

Customers were categorized as those who took precision supplements (PS) along with personalized food recommendations or those who did not (controls). Eligible PS subjects were defined as those having a continuous monthly PS subscription from the purchase of a kit (t0) to the second kit (t1). Subjects (both PS and controls) were only included if they were naive to PS, meaning no previous subscription before t0. Subjects were only included if they demonstrated symptoms of depression at the time of purchasing the first kit. Subjects were screened for depression with the patient health questionnaire 2 (PHQ-2) and their symptoms were evaluated with the patient health questionnaire 9 (PHQ-9) [12]. Participants in the control group who followed Viome food recommendations in the Viome app were excluded.

### 2.2. Precision supplements

Viome’s precision supplements result from combining microbiome and human activity profiling using metatranscriptomics (gene expression) and an AI-powered recommendation engine. The former identifies the active microorganisms and human molecular functions based on which a set of Viome wellness scores are computed to reflect the individual’s gut health [11]. The latter implements a precision supplement recommendation algorithm that takes as input the Viome wellness scores and self-reported phenotypes for an individual and produces as output a set of ingredients with dosages targeting all Viome-assigned conditions for that individual. The algorithm is a multi-dimensional constrained optimization reasoning engine that takes into account a wide variety of factors and decides the optimal set of ingredients for a given individual, out of hundreds of available ingredients [11]. PS can include any type of supplementation plan including precision prebiotics and probiotics, precision supplements like vitamins and herbal extracts, or a combination of both. The Viome Precision Supplement manufacturing facility then turns those ingredients and dosages into appropriate packets and powders to be delivered to that individual.

It is important to note that each individual in the PS group in this study received precision supplements that targeted the entire collection of health conditions used by the algorithm, rather than only one specific condition, such as depression. As an example, a person could present depression symptoms, self-reported type 2 diabetes, and have an allergy to nuts. In that case, the precision supplements will target the three conditions at once.

### 2.3. Patient Health Questionnaire

The primary endpoint was the Patient Health Questionnaire-9 (PHQ-9) score. PHQ-9 is an instrument for making criteria-based diagnoses of depression and other mental disorders commonly encountered in primary care [12]. It contains 9 items and consists of the actual 9 criteria upon which the diagnosis of DSM-V depressive disorders is based. The first two questions of PHQ-9 comprise PHQ-2, which is a screening tool. Classification of the PHQ-9 is “minimal” (total score 0–4), “mild” (total score 5–9), “moderate” (total score 10–14), “moderately severe” (total score 15-19) or “severe” (total score 20-27).

Similar to other studies [12], for this analysis, we only included those participants with moderate, moderately severe, and severe symptoms (score >= 10) at t0. Participants who scored between 0 and 9 on the PHQ-9 questionnaire at t1 were included and categorized as having no/low depression.

### 2.4. Statistical analysis

We evaluated categorical changes across depression categories from t0 to t1 using the Mann -Whitney U test.

To evaluate the effect of PS on depression, we used multiple group propensity score weighting in the context of parametric difference in differences (DD) models [14], where observations were weighted to ensure similarity in age, sex, and physical activity, measured by the international physical activity questionnaire (iPAQ), and body mass index (BMI). Missing values for BMI and physical activity were imputed. Adjusted p-values were calculated to account for Type I errors.

Descriptive analyses were used to report baseline characteristics, including depression medication use, and time between kits in the study population. All analyses were implemented using Python 3.6.5. P-values are reported and interpreted appropriately in context.

## 3. Results

A total of 86 PS and 45 controls responded to the PHQ-9 questionnaire at t0, and 60 and 29 at t0 and t1 (Figure 1). Baseline characteristics were generally well-balanced between the two groups (Table 1).

**Table 1.** Descriptive characteristics at t0

|  | PS (n=86) | Control (n=45) | p-value |
| --- | --- | --- | --- |
| Age (mean +/- SD) | 46.5 +/- 13.0 | 42.9 +/- 15.0 | 0.15 |
| Female (mean +/- SD) | 60 | 51.7 | 0.46 |
| BMI (mean +/- SD) | 26.3 +/- 6.1 | 27.3 +/- 7.7 | 0.39 |
| iPAQ (mean +/- SD) | 1.0 +/- 0.9 | 1.2 +/- 0.8 | 0.21 |
| t1-t0 (days, mean +/- SD) | 192.9 +/- 58.9 | 189.5 +/- 90.2 | 0.83 |
| Under depression medication | 22 | 21 | 0.92 |

BMI, body mass index; iPAQ: international physical activity questionnaire

First, we consider a categorical view of the PHQ-9 score. Starting at t0, the proportions of moderate, moderately severe, and severe people were similar in the control and PS group (Table 2). At t1, clinically significant categorical changes between controls and PS were observed (Figure 2; effect size = 0.48; p <0.001). Overall, 60 out of 86 (69.8%) in the PS group improved their PHQ-9 category from moderate, moderately severe, and severe to no/low depression versus only 7 out of 45 participants (15.6%) in the control group (Figure 3). Additionally, 67 out of 86 (77.9%) in the PS group improved their PHQ-9 category by at least one category, versus only 10 out of 45 (22.2%) in the control group.

**Table 2.**
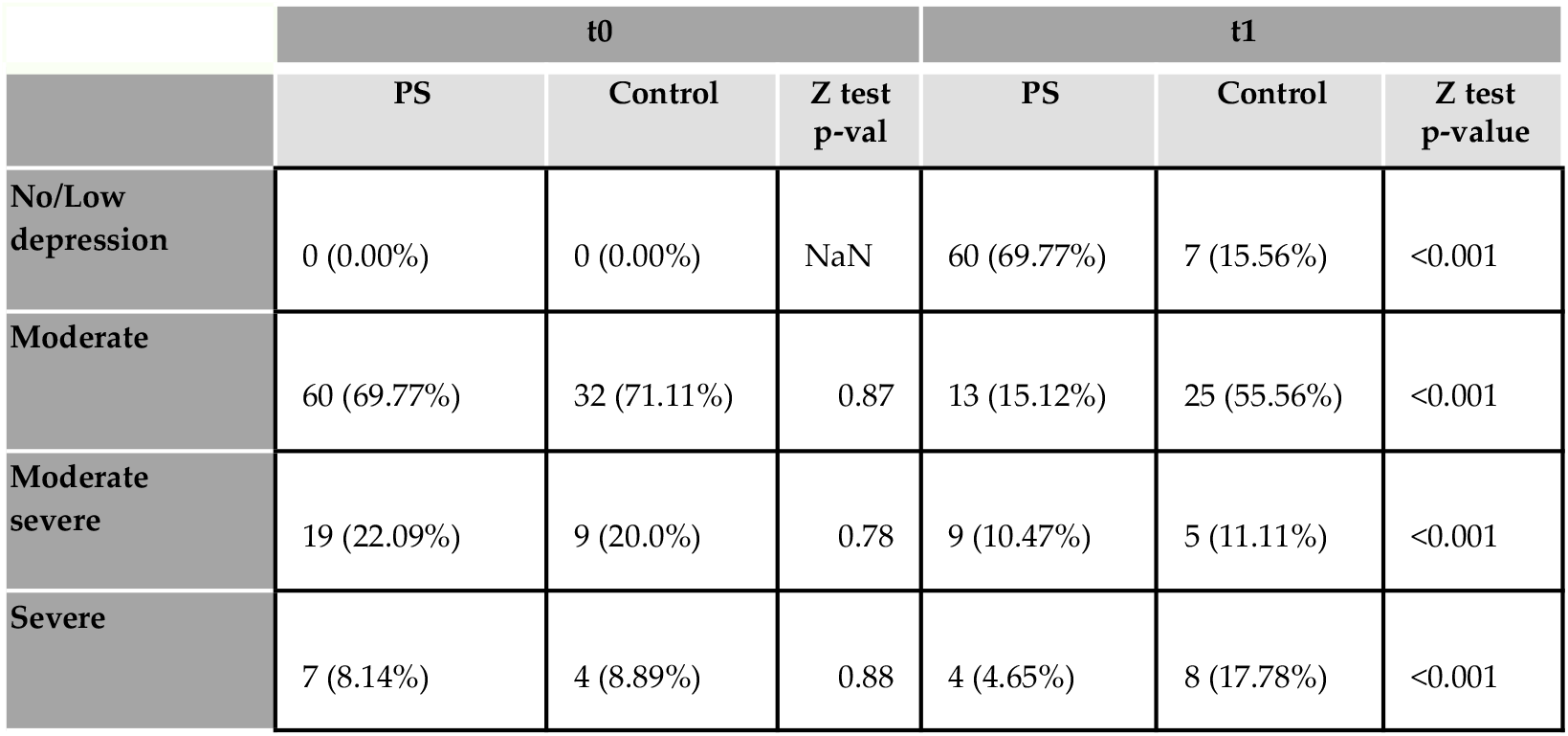
PHQ-9 depression category changes

**Figure 2.**
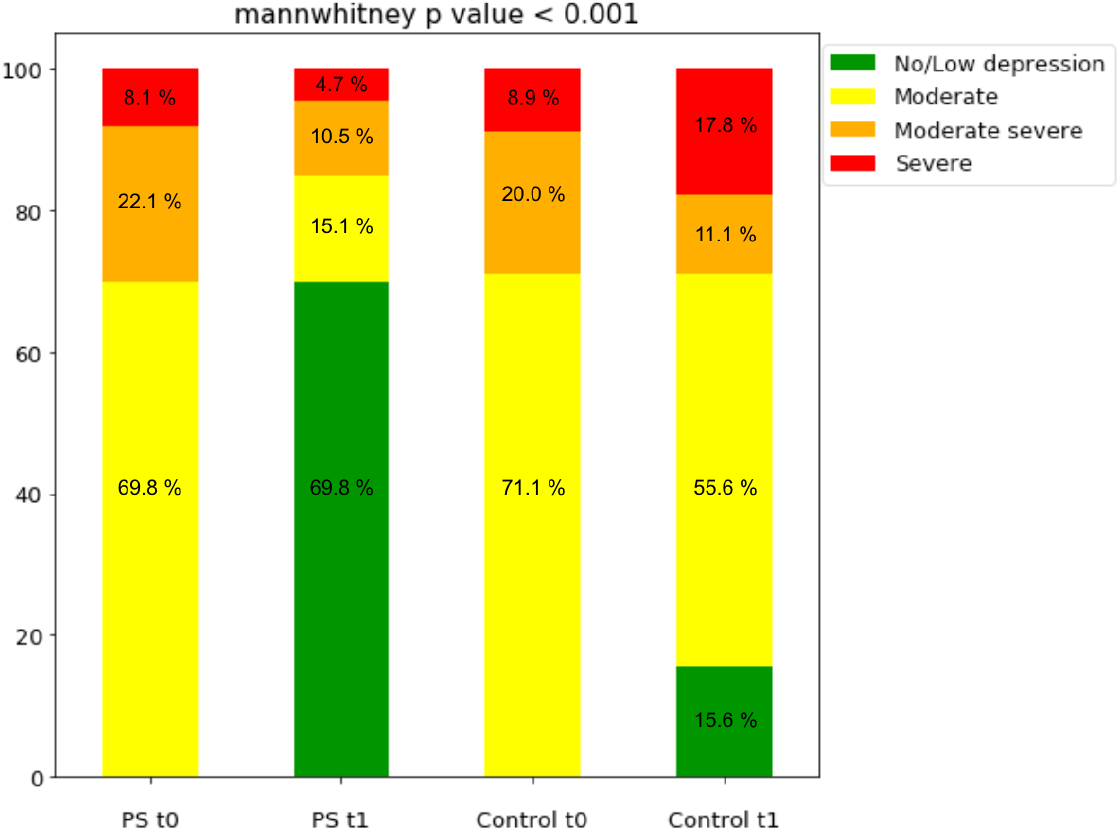
Depression categorical changes from t0 to t1

**Figure 3.**
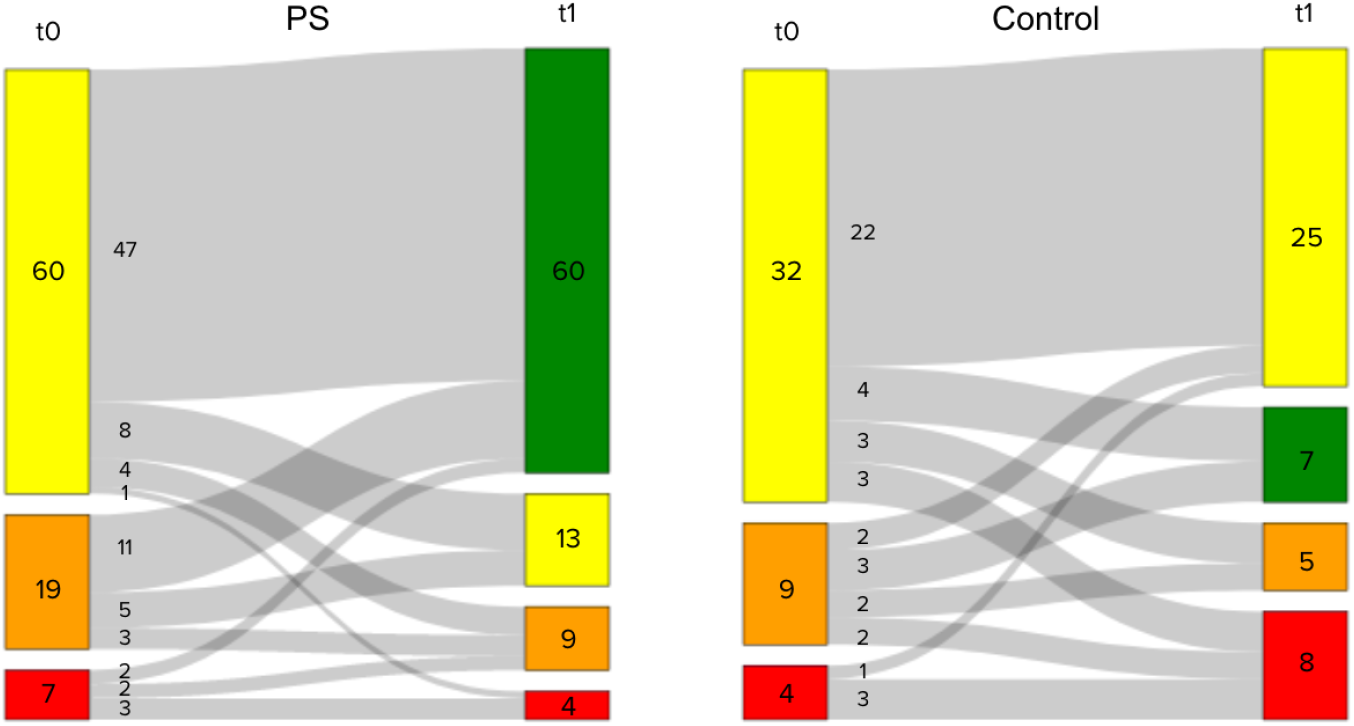
Patients’ journey of PHQ-9 categories from t0 to t1

Next, we consider a continuous view of the PHQ-9 score. At t0, the PHQ-9 scoring average in both cohorts was moderate. At t1, a higher number of PS people improved their score (∼4 points decrease) compared to controls (Table 3). DD was observed between PS and controls when adjusting for age, sex, physical activity, and BMI, although the analysis was underpowered to detect a clinically significant difference (DD: -2.11, p-value = 0.22) (Figure 4).

**Table 3.** PHQ-9 score changes

|  | PS | Control |
| --- | --- | --- |
| t0 | 13.75 +/- 3.80 | 14.07 +/- 3.64 |
| t1 | 9.78 +/- 6.42 | 13.59 +/- 6.65 |

**Figure 4.**
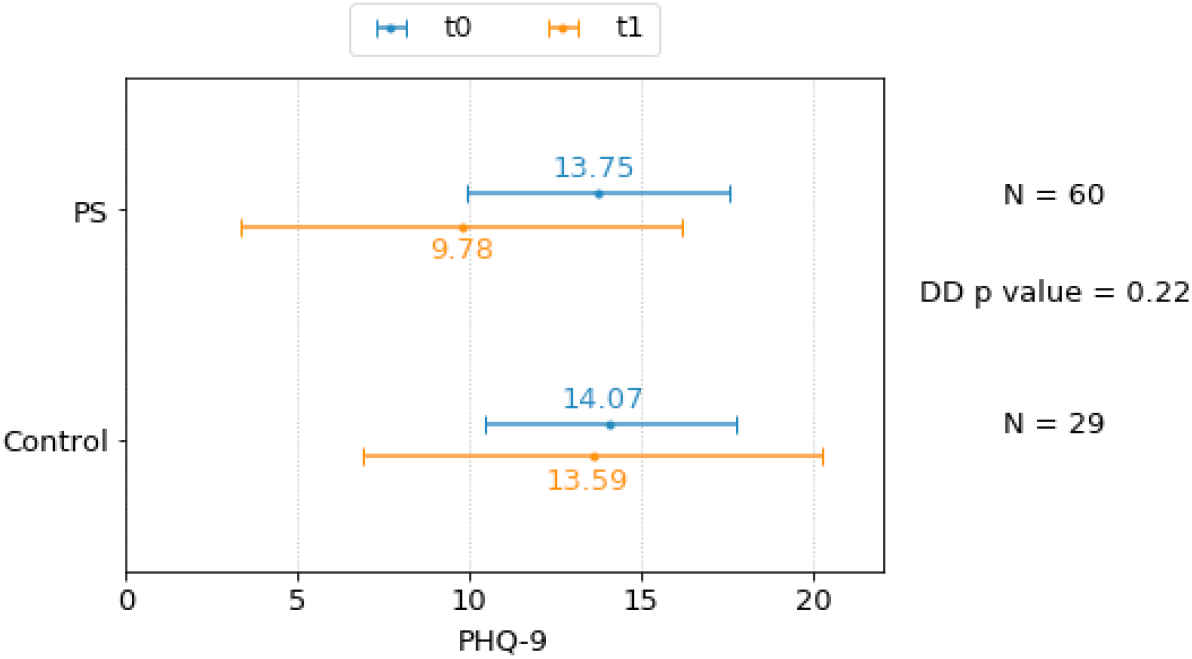
PHQ-9 depression score changes from t0 to t1

## 4. Discussion

To our knowledge, this is the first study evaluating the effectiveness of precision supplements in depression symptoms as part of a personalized nutrition subscription plan that takes into account the gene expression of the gut microbiome and human cells and self-reported phenotype conditions to improve an individual’s overall health. Population-level studies have repeatedly demonstrated a link between aspects of nutrition and depression, although most of the time the effect sizes were not clinically significant [15]. In this study, depression symptoms measured by PHQ-9 decreased by more than 4 points among people who followed Viome’s precision supplements for an average of ∼6 months, and clinically significant differences between people who followed Viome’s precision supplements versus those who did not were observed. A higher proportion of participants subscribed to the PS program improved their depression symptoms (as measured by PHQ-9 categories) compared to those who did not follow the Viome food recommendations (77.3% vs. 20.8%).

Some studies have evaluated the effects of combining vitamins and nutrient supplementation as depression treatments in different populations [16,17] including pregnant women [18] and people with obesity [19]. Nguyen et al. [18] reported that the intervention of multiple micronutrients and iron-folic acid groups had a significant impact on lowering depression scores during the first and second trimesters of pregnancy compared to folic acid alone. In contrast, among overweight or obese adults, multinutrient supplementation including omega-3 PUFAs, vitamin D, folic acid, and selenium compared with placebo did not reduce episodes of major depressive disorder during 1 year [19]. While these studies focused on a specific target population, the precision supplements included in our study were personalized according to the phenotype condition and microbiome activity of each individual.

Selected ingredients from an array of more than 100 active compounds, food extracts, and dietary supplements were recommended for each of the participants taking the precision supplements. Personalized supplements have the advantage of considering individual preferences, allergies, and health conditions. US adults with depression often experience other comorbidities [1]. Between 31–45% of patients with coronary artery disease (CAD), including those with stable CAD, unstable angina, or myocardial infarction (MI), suffer from clinically significant depressive symptoms [20]. In our sample, 14.7% of participants presented at least one other health condition (data not shown). A holistic approach where supplements are offered based on individual circumstances seems to be effective in reducing depression symptoms.

Our study has some limitations inherent to the study design. This observational study included adults who purchased precision supplements, so the intervention could not be randomized. The control group was naive to supplements and food recommendations but we cannot know if they were taking other dietary supplements to treat depression. Similarly, we only know the PS group was receiving the supplements, but we cannot track supplement intake. We do not know if participants followed psychological counseling or therapy at the time of the study. We could evaluate, though, the number of people taking medication to treat depression and we observed it was similar in both groups. Lastly, one could argue that the control group purchased a second kit when they were not feeling well as opposed to doing it as part of a subscription plan like the PS did. While this is plausible, the average time between the two purchased kits was similar in both groups (^∼^192 days) which makes us think this is unlikely.

## 5. Conclusions

After ∼6 months of use, precision supplements delivered by an AI-driven recommendation system based on gene expression showed a significant beneficial effect on individuals with depression symptoms, measured with the clinically validated questionnaire PHQ-9. 69.8% of the individuals in the intervention group improved their symptoms compared to 15.6% in the control group. A follow-up placebo-controlled randomized control trial that also evaluates the specific role of supplements on the host microbiome of people with depression is already in progress.

## Data Availability

The data used in this study, beyond what has been summarized in this paper, cannot be shared publicly due to privacy and legal reasons. Researchers requiring more data for non-commercial purposes can request more data via our data access website: https://www.viomelifesciences.com/data-access. Upon review, Viome may provide access to more summary statistics through a Data Transfer Agreement that protects the privacy of our participants' data.

## Author Contributions

Conceptualization, CJ, NS, MM, and GB; methodology, CJ, NS, MM, GA, JC, HK, and GB; software, NS, MM, VG, AG, and EP; formal analysis, CJ, NS, and MM; resources, MM, and GB; data curation, NS, LH, VG, AG, and EP; writing—original draft preparation, CJ, NS, MM, LH, and GB; writing—review and editing, all authors; visualization, CJ, and NS; supervision, CJ, LH, and GB; project administration, CJ; funding acquisition, MV, and GB. All authors have read and agreed to the published version of the manuscript.

## Informed Consent Statement

Informed consent was obtained from all subjects involved in the study.

## Ethics statement

The Viome ethics process assessed the research project to determine whether it needed to be reviewed by the Viome IRB. Based on the US Health and Human Services CFR 46.104, section 4(II), it was determined that the research in “The effectiveness of precision supplements on depression symptoms in a US population” is exempt from IRB reviews and oversight due to: (a) this research uses data from Viome customers who have consented to the terms in https://www.viome.com/terms (see esp. clause 26) (b) the research as contained in solely uses retrospective de-identified Viome customer data, and (c) the research data analysts do not contact the customers or re-identify them.

